# Endoscopic findings in patients with persistent dyspepsia in a tertiary care hospital of south Punjab

**DOI:** 10.1101/2021.12.23.21268300

**Authors:** Farooq Mohyud Din Chaudhary, Muhammad Asif Gul, Rizwan Hameed, Yasir Abbas Zaidi, Shehryar Kanju, Muhammad Ilyas, Muhammad Zubair, Qasim Umar, Syeda Manal Altaf, Ahsan Tameez-ud-din

## Abstract

**Background:** One of the most common clinical problem encountered by physicians in clinical practice is dyspepsia. This symptom has great impact on quality of life of patients. There are numerous causes of dyspepsia, organic as well as functional. Endoscopy is the diagnostic test of choice in these patients.

**Aim:** The aim of our study was to see the endoscopic findings in patients with persistent dyspepsia.

**Methods:** Retrospective analysis of data of patients who underwent Esophagogastroduodenoscopy (EGD) for persistent dyspepsia was collected and evaluated.

**Results:** There were 495 patients in our study, 244 females and 251 males, with a mean age of 41 years. Almost half of the patients belonged to 21-40 years age group. The most common endoscopic finding in patients with persistent dyspepsia was gastritis (n=219, 44.2%), followed by normal endoscopy (n= 94, 19%), incompetent lower esophageal sphincter (n=67, 13.5%), gastric malignancy (n=48, 9.7%). Ulcer disease was found in just 15 patients (3%).

**Conclusion:** Most common endoscopic finding in patients with persistent dyspepsia was gastritis followed by normal endoscopy.

## INTRODUCTION

Dyspepsia is defined by the presence of heartburn and/or abdominal pain or discomfort located in the upper abdomen. Approximately one third of adults experience dyspepsia annually [1]. Substantial cost is involved in the evaluation, and treatment of dyspepsia, especially work loss [2]. The American Gastroenterological Association (AGA) and American Society for Gastrointestinal Endoscopy (ASGE) have published guidelines for the role of endoscopy in select number of patients who have dyspepsia. These organizations recommend EGD for patients with dyspepsia who are more than 50 years of age and have alarm symptoms such as dysphagia, weight loss, gastrointestinal bleeding and symptoms refractory to appropriate empiric medical therapy such as proton pump inhibitors [3,4].

The role of endoscopy in young patients with dyspepsia without alarm symptoms is controversial as a lot of times the cause is functional in nature. However many a times endoscopy becomes mandatory in these young patients who have symptoms of persistent dyspepsia refractory to medical treatment. The causes of persistent dyspepsia range from a wide variety of functional and organic disorders including peptic ulcer disease, tumors of the esophagus and stomach and sometimes rare causes which include pancreatic rest and duodenal diverticuli [5, 6].

The aim of our study was to evaluate patients who had persistent dyspepsia despite taking proton pump inhibitors. These patients were evaluated with EGD to see the endoscopic findings.

## METHODS

This retrospective analysis of endoscopic findings in patients with persistent dyspepsia was conducted at the department of gastroenterology Nishtar hospital Multan. Records from the endoscopy register were extracted. Data was collected from 2018 to 2020. All patients of both genders who underwent Esophagogastroduodenoscopy (EGD) for symptoms of persistent dyspepsia despite proton pump inhibitors therapy were included in this study. Pregnant ladies, patients with malignancy of any kind were excluded from the study. Due to the retrospective nature if this study, ethical review was waived off for this study.

From the endoscopy register patient’s age, gender, indications and findings on EGD were recorded on a pre-designed Performa. Confidentiality of the patients was ensured. The collected data was entered and evaluated in Statistical Package for Social Sciences (SPSS) version 20 (IBM Corp, Armonk, US). The results were reported as frequencies, percentages, and tables.

## RESULTS

There were 495 patients who fulfilled the inclusion criteria. The mean age of study population was 41 years with a standard deviation of 16 years. There were 244 females (49.3%) and 251 males (50.7%) in our study. The youngest person in our study was 13 years of age, while the eldest was 89 years of age.

Table 1 shows the age wise distribution of our study population who underwent EGD. Almost half of the patients belonged to the 21-40 years age group. Table 2 shows the endoscopic findings of patients with persistent dyspepsia. The most common finding in these patients was gastritis, seen in 219 patients (44.2%). Normal endoscopy was seen in 94 patients (19%), being the second most common finding in these patients. Incompetent lower esophageal sphincter was seen in 67 patients (13.5%). Gastric malignancy was seen in 48 patients (9.7%). Other less common findings are shown in Table 2.

**Table 1:** Age wise distribution of study population (N=495).

| **Age Group (Years)** | **Frequency (n)** | **Percent (%)** |
| --- | --- | --- |
| <=20 | 41 | 8.3 |
| 21-40 | 237 | 47.9 |
| 41-60 | 159 | 32.1 |
| 61-80 | 54 | 10.9 |
| >80 | 4 | 0.8 |

**Table 2:**
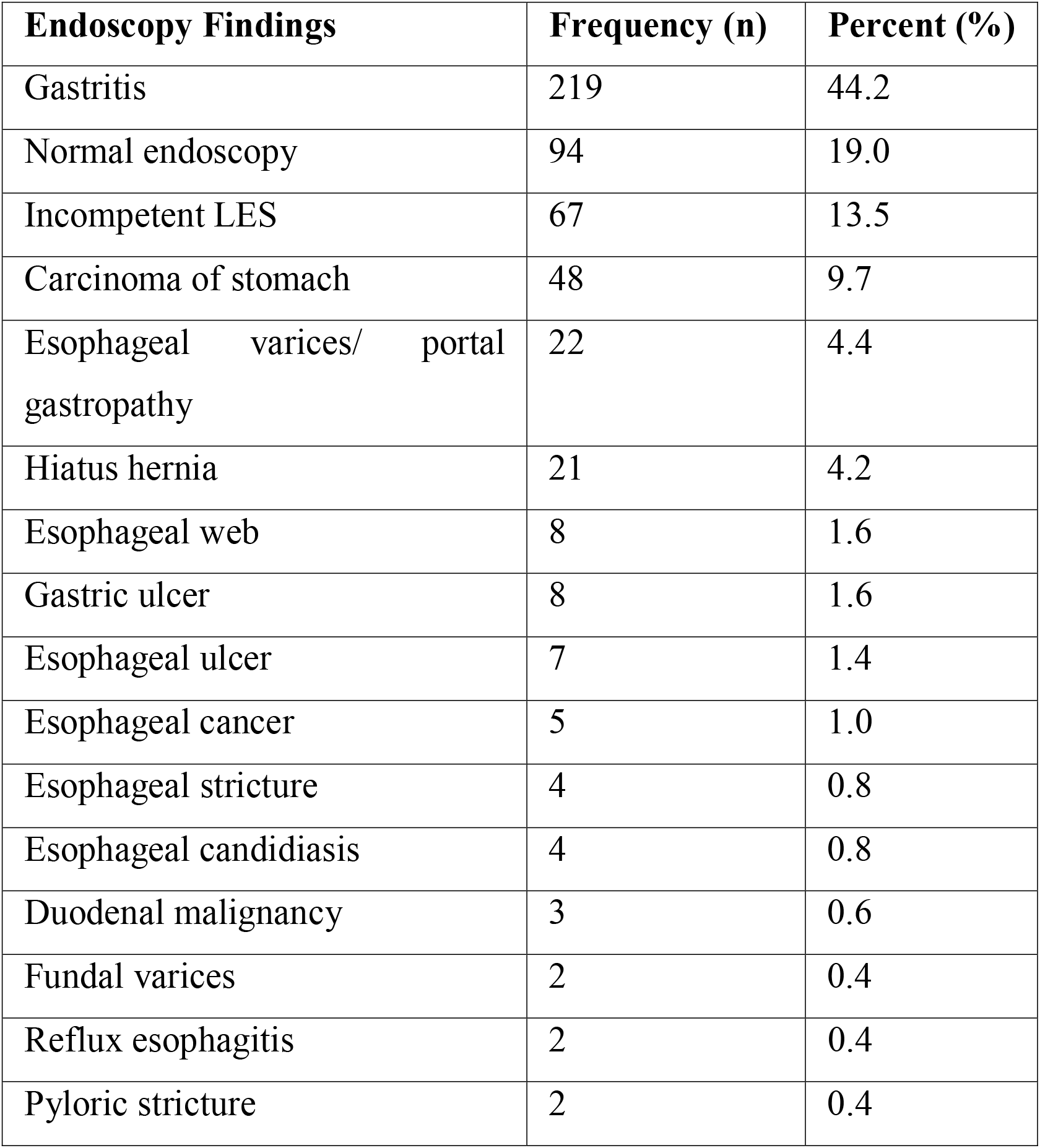
Endoscopy findings of patients with persistent dyspepsia (N=495)

## DISCUSSION

Our study is an attempt to update the existing clinical data regarding the endoscopic findings in patients of persistent dyspepsia especially in this part of the world. Our study shows that more than half of the study population was younger than 40 years of age This was in contrast to an international study where 36% patients were younger than 50 years of age [7]. Our study shows that the most common endoscopic finding in patients with persistent dyspepsia is gastritis (44%). Other studies from around the world found gastritis in 11% [8], 26% [9] and in one study erosive gastritis was found very infrequently [10].

Normal endoscopy was found in 19% of our patients. However studies around the world showed a higher prevalence of normal endoscopy ranging from 26% to 50% [8-10]. This might be due to stringent criteria/alarm symptoms for short listing patient for endoscopy in this institute.

Malignancy of the stomach was found in almost 10 percent of our study, which was much higher as compared to other studies which revealed a cancer rate of 1-3% [8-10]. This might be due to late presentation of patients to a physician for evaluation, stigma of endoscopy in general public and lack of availability of endoscopic services in many areas.

Infrequent endoscopic findings in our study included varices, portal gastropathy, web and stricture. Esophageal and fundal varices which mostly are the cause of upper gastrointestinal bleeding [11, 12], were found in 4.4% and 0.4% of patients in our study, respectively. This might be due to the high prevalence of hepatitis B and C in this region leading to cirrhosis and portal hypertension.

Endoscopy for dyspepsia is done infrequently, 15% in a local study [13]. However, it is imperative that whenever alarm symptoms are observed, EGD should be done in patients to evaluate for more sinister symptoms.

## CONCLUSION

Persistent dyspepsia should always be evaluated with endoscopy. Our study updates existing data regarding endoscopic findings in patients with persistent dyspepsia. We found that gastritis was the most common finding, followed by normal endoscopy, incompetent LES and gastric malignancy. Ulcer disease was rare in our study.

## Data Availability

All data produced in the present study are available upon reasonable request to the authors

## Notes

**Conflict of interest statement:** The authors have no conflict of interest to disclose.

### Competing Interest Statement

The authors have declared no competing interest.

### Funding Statement

This study did not receive any funding

### Author Declarations

This study was approved by the Institutional Review Board of Nishtar Medical University

## REFERNCES

1. Barbara L, Camilleri M, Corninaldesi R. Definition and investigation of dyspepsia: consensus of an International Ad Hoc Working Party. Dig Dis Sci 1989;34:1271–1276.

2. Sandler RS, Everhart JE, Donowitz M, et al. The burden of selected digestive disease in the United States. Gastroenterology 2002;122:1500–1511

3. Eisen GM and the Standards of Practice Committee. The role of endoscopy in dyspepsia. Gastrointest Endosc 2001;54:815–817.

4. Talley NJ, Silverstein MD, Agreus L, et al. AGA technical review: evaluation of dyspepsia. Gastsroenterology 1998;114:582–595.

5. Siddiqui KH, Tameez Ud Din A, Chaudhary FMD, et al. S3602 Pancreatic Rest Causing Unrest, The American Journal of Gastroenterology: October 2020 - Volume 115 - Issue - p S1860 doi: 10.14309/01.ajg.0000716456.74020.8f

6. Tameez Ud Din A, Siddiqui KH, Chaudhary FMD, et al. S3574 Duodenal Diverticula: An Unusual Cause of Dyspepsia, The American Journal of Gastroenterology: October 2020 - Volume 115 - Issue - p S1845 doi: 10.14309/01.ajg.0000716344.13446.06

7. Lieberman D, Fennerty MB, Morris CD, et al. Endoscopic evaluation of patients with dyspepsia: results from the national endoscopic data repository. Gastroenterology. 2004 Oct;127(4):1067–75. doi: 10.1053/j.gastro.2004.07.060. PMID: 15480985.

8. Andrabi RUS, Ahad WA, Yousuff M, et al. Endoscopic Findings in Persistent Dyspepsia in Secondary Care Hospital Setting in North Kashmir. J Assoc Physicians India. 2019 Sep;67(9):46–49. PMID: 31561689.

9. Kaosombatwattana U, Charatcharoenwitthaya P, Pausawasdi N, et al. Value of age and alarm features for predicting upper gastrointestinal malignancy in patients with dyspepsia: an endoscopic database review of 4664 patients in Thailand. BMJ Open. 2021 Oct 27;11(10):e052522. doi: 10.1136/bmjopen-2021-052522. PMID: 34706958; PMCID: PMC8552171.

10. Heikkinen M, Pikkarainen P, Takala J, et al. Etiology of dyspepsia: four hundred unselected consecutive patients in general practice. Scand J Gastroenterol. 1995 Jun;30(6):519–23. doi: 10.3109/00365529509089783. PMID: 7569757.

11. Malghani WS, Malik R, Chaudhary FMD, et al. Spectrum of Endoscopic Findings in Patients of Upper Gastrointestinal Bleeding at a Tertiary Care Hospital. Cureus 11(4): e4562. DOI 10.7759/cureus.4562

12. Malghani WS, Chaudhary FMD, Tameez ud Din A, et al. Frequency of fundal varices in cirrhotic patients presenting with upper gastrointestinal bleed. Professional Med J 2019; 26(10):1655–1659. DOI: 10.29309/TPMJ/2019.26.10.3164

13. Chaudhary FMD, Gul MA, Hameed R, et al. Three year experience of Upper Gastrointestinal Endoscopic procedures at a tertiary care hospital of South Punjab. medRxiv 2021.11.11.21266215; doi: https://doi.org/10.1101/2021.11.11.21266215

